# Remodeling of α1-Acid Glycoprotein Glycoforms During Long-Term Calorie Restriction

**DOI:** 10.64898/2026.10.30.26366848

**Authors:** Divya Maikhuri, Mary Thompson, Tea Pribić, Helena Deriš, Jelena Šimunović, Siniša Habazin, Jerko Štambuk, Azra Frkatović-Hodžić, Kim M Huffman, William E Kraus, Sai Krupa Das, Gordan Lauc, Sridevi Krishnan

**Affiliations:** School of Nutritional Sciences and Wellness, The University of Arizona; College of Nursing, The University of Arizona; Genos Ltd, Glycoscience Research Laboratory, Zagreb, Croatia; Duke Molecular Physiology Institute and Department of Medicine, Duke University School of Medicine, Durham, North Carolina, USA; Jean Mayer, USDA, Human Nutrition Research Center on Aging at Tufts University, Boston, Massachusetts, USA

**Author notes:** Corresponding author contact info: Sridevi Krishnan, PhD.

**Keywords:** calorie restriction, glycomic biomarkers

## Abstract

AGP1 (α-1 acid glycoprotein) is an acute phase immune response protein recently shown to influence leptin receptor activity and body weight regulation. AGP is a highly glycosylated protein, with 45% of its molecular weight made up by glycans, yet little is known about how the latter responds to metabolic interventions or its role in AGPs immunometabolic function. In this pilot study (n = 26), we evaluated the effect of calorie restriction (CR - 2years, 25% CR in non-obese adults) on N-glycosylation of AGP1. AGP1 has 5 glycosylation sites, of which sites IV and V are dominated by highly branched, fucosylated, and sialylated glycans, and inflammatory conditions are associated with reduced branching at sites IV and V and increased fucosylation. With CR (0 month to 24 month, ∼12% CR, weight loss of ∼8.2 kg), there was increase in branching at sites IV and V (p<0.05, FDR corrected), and increased fucosylation (p<0.05, FDR corrected). Exploratory moderation analyses suggest that highly sialylated site IV glycans and low-branched site I glycans may influence the relationship between CR and leptin (p<0.05). Overall, AGP glycome is largely remodeled with CR, and it is unknown if these are upstream or downstream of immunometabolic changes seen with CR.

## Introduction

Alpha-1-acid glycoprotein (AGP, also known as orosomucoid, ORM) is an acute-phase protein whose expression increases markedly in response to inflammatory stimuli [1-3]. While the liver is the principal source of circulating AGP, additional expression occurs in adipose tissue, vascular endothelium, immune cells, and the nervous system [3-5]. AGP contributes to host defense by modulating immune responses, including reducing lymphocyte proliferation, impairing neutrophil chemotaxis, limiting superoxide production, and inhibiting platelet aggregation [6,7]. Beyond its immune roles, AGP is increasingly recognized as a regulator of metabolic processes. Circulating AGP levels are positively associated with adiposity, serum leptin, and fasting glucose in humans [8], and in murine models, AGP has been proposed as a leptin receptor agonist capable of dampening leptin signaling [9]. This dual function suggests that AGP may serve as an immunometabolism marker integrating inflammatory cues with pathways regulating energy balance.

AGP is among the most extensively glycosylated proteins in human plasma, with carbohydrates contributing nearly half of its molecular mass, primarily in the form of sialylated N-linked glycans [10]. The protein contains five conserved N-glycosylation sites (Asn-33, -56, -72, -93, -103 – sites I - V) that can accommodate glycans with diverse branching patterns and terminal modifications, generating substantial structural heterogeneity [10,11]. Specifically, sites I and II (Asn33 and Asn56) appear to have predominantly fully sialylated tri-antennary glycans, while sites III-V (Asn72, Asn93, and especially Asn103) are enriched in more highly branched tetra-antennary and antennary-fucosylated structures. Also, this glycan diversity is not static. Glycosylation patterns shift in response to physiological and pathological state changes, and this shift is different by site[12]. Notably, increases in α1,3-fucosylation and increase in bi-antennary structures, specifically at sites III-V are reported in conditions such as sepsis, rheumatoid arthritis, cancer, and diabetes [12-15]. On the other hand, higher order branching (tetra-antennary species) and reduction in 1,3 fucosylations have been noted in pregnancy and estrogen treatment[12,15]. Functional studies suggest that AGP’s biological effects are dictated largely by its glycan composition rather than its polypeptide core [16]. Since N-glycosylation is altered in different disease states, studying glycoform-specific changes offers critical insight into how AGP contributes to both immune regulation and metabolic control, and its potential as a biomarker of systemic physiological change [17,18].

The Comprehensive Assessment of Long-Term Effects of Reducing Intake of Energy (CALERIE 2) was a two-year randomized controlled trial that evaluated sustained 25% Calorie Restriction (CR) in non-obese humans, with primary outcomes centered on resting metabolic rate and core temperature, and secondary outcomes including circulating biomarkers of health and metabolism [19]. A recent pilot analysis of plasma, IgG and complement C3 N-glycome in a subset of 26 CALERIE2 participants indicated that CR might increase galactosylation of IgG and reduce branching of plasma protein N-glycans [20]. These findings suggest that long-term CR modifies protein N-glycosylation in ways that may reflect systemic adaptations in metabolism and immune function. Building on this work, we present a preliminary investigation focusing specifically on AGP glycosylation in the same participant subset. Here, we characterize longitudinal changes in AGP glycosylation at baseline, 12 months, and 24 months of CR, and examine whether AGP glycoform changes are associated with or capable of predicting changes in metabolic, energy balance, and satiety markers.

## Methods

### Study Design

The present study utilized data from the Comprehensive Assessment of Long-term Effects of Reducing Intake of Energy Phase 2 (CALERIE 2) trial, a randomized controlled intervention conducted across multiple clinical sites [21]. Detailed descriptions of the trial rationale, design, and intervention protocols have been published previously, and full documentation is publicly available through the Aging Research Biobank (https://agingresearchbiobank.nia.nih.gov/). The CALERIE 2 protocol (ClinicalTrials.gov identifier: NCT00427193; registered January 25, 2007) was approved by the Institutional Review Boards at Washington University School of Medicine (St. Louis, MO), Pennington Biomedical Research Center (Baton Rouge, LA), and Tufts University (Boston, MA), as well as by the coordinating center at Duke University (Durham, NC). All participants provided written informed consent prior to enrollment.

### Participants

CALERIE 2 enrolled healthy adults who were randomly assigned in a 2:1 ratio to either a calorie restriction (CR) intervention or an ad libitum (AL) control condition. A total of 220 individuals were randomized; after early withdrawals, 143 participants initiated the CR intervention, and 75 participants comprised the AL control group. The intervention period lasted 24 months. Participants were recruited from the community and were eligible if they were healthy men aged 21–50 years or premenopausal women aged 21–47 years with a body mass index (BMI) between 22.0 and <28.0 kg/m². Individuals were excluded if they had cardiometabolic disease, abnormal clinical laboratory values, psychiatric or behavioral conditions, or were using medications known to affect metabolism (with the exception of oral contraceptives). To minimize confounding from high or changing activity levels, individuals engaging in high volumes of structured physical activity were also excluded. Participants assigned to the calorie restriction group were prescribed an immediate and sustained reduction in energy intake targeting a 25% decrease from individual baseline energy requirements. Baseline energy needs were determined using the intake-balance method, with total daily energy expenditure objectively measured via doubly labeled water over two consecutive 14-day periods [22].

For this pilot analysis, a subset of 26 participants was selected from the CR intervention arm. Inclusion in this subsample was limited to individuals who completed the full 24-month trial and demonstrated the greatest adherence to the prescribed intervention, achieving average (over 24 months) calorie restriction of 12%. This selection strategy was implemented to maximize contrast in biological responses associated with sustained energy restriction.

### Data access

Plasma samples and associated clinical data from the CALERIE 2 trial were accessed following approval from both the CALERIE Steering Committee and the National Institute on Aging (NIA) Aging Research Biobank. All samples were de-identified and re-coded to ensure that personnel conducting assays were blinded to participant identity and study time point. Plasma samples were obtained at baseline, 12 months, and 24 months and are maintained by the NIA Aging Research Biobank, where they are available to qualified investigators upon request. In addition to biospecimens, comprehensive phenotypic and clinical data were retrieved from the CALERIE Biorepository. Variables obtained for the present analyses included demographic and anthropometric measures (age, body mass index, body weight, and body composition), indices of energy intake, total daily energy expenditure, and calculated percent calorie restriction, immune and inflammatory markers assessed in plasma included interleukin-1B (IL-1B), interleukin-6 (IL-6), interleukin-8 (IL-8), and C-reactive protein (CRP), clinical and metabolic variables included plasma concentrations of high-density lipoprotein cholesterol, low-density lipoprotein cholesterol, triglycerides, and total cholesterol, as well as cortisol, fasting glucose, fasting insulin and oral glucose tolerance test postprandial glucose and insulin. Derived indices of insulin sensitivity and β-cell function, Homeostasis Model Assessment of Insulin Resistance (HOMA-IR) and Homeostasis Model Assessment of β-cell function (HOMA-β), were also included.

### AGP N-glycome quantification

Samples were processed following our established protocol [23,24]. Twenty microliters of plasma were mixed with 80 µL of 0.75 M perchloric acid and centrifuged; the supernatant was retained. Phosphotungstic acid was then added to precipitate the “seromucoid” fraction, which was collected after centrifugation and the supernatant discarded. The precipitate was solubilized with NaOH. The solubilized material was treated with 2.5 µL of 1.5% RapiGest to aid denaturation and subsequent digestion. Disulfide bonds were reduced with dithiothreitol and alkylated with iodoacetamide. The pH was adjusted with ammonium bicarbonate. Samples were digested overnight with 0.8 µg of trypsin. RapiGest was degraded by adding HCl, and the resulting glycopeptides were enriched by binding to preconditioned HILIC beads, washed, and eluted in ultrapure water containing 0.1% TFA. Eluates were dried in a vacuum concentrator and stored at 20 °C until analysis. Dried glycopeptides were reconstituted in ultrapure water and separated on a nanoACQUITY UPLC system (Waters) coupled to a Compact mass spectrometer (Bruker Daltonics). Automated quantification of the acquired MS data was performed with LaCyTools (version 1.1.). After QC, absolute signal intensities were normalized to the totalerea for each glycosylation site.

#### AGP nomenclature

Human AGP has five N glycosylation sites at Asn-33, -56, -72, -93, -103 on the polypeptide backbone [25], and are referred to as sites I – V (**Supplementary Figure 1**). Similarly, in our dataset, AGP glycopeptides are labeled to indicate both N-glycosylation site and AGP isoform/allele. IORM, IIORM, IIIORM, IVORM and VORM denote glycopeptides arising from N-glycosylation sites I, II, III, IV and V respectively (as stated earlier), on orosomucoid, without further isoform specification. ORM1 and ORM2 refer to AGP1 and AGP2 respectively. The combined label ORMISORMIIA denotes a peptide sequence shared between specific allelic variants of AGP1 and AGP2 (ORMIS and ORMIIA) that cannot be distinguished by the MS workflow and is therefore treated as a joint AGP1/AGP2 glycopeptide species. Also, within this framework, IORMIF designates a site I glycopeptide compatible with F variant ORM sequences (e.g.: ORM1*FType), again reflecting a form that is not uniquely resolvable to a single isoform but instead aggregates F allele–related site I glycoforms across ORM1/ORM2 [26,27]. Further, the nomenclature specifies the glycan composition using standard abbreviations, where the subscript x indicates the number of residues of each monosaccharide type (N = N-acetylhexosamines, H = Hexose, F = Fucose, S = N-acetylneuraminic acid).

Relative intensities of individual AGP glycoforms were measured in CALERIE 2 using LC-MS and used as the foundation for all analyses in this study. Site-derived AGP glycosylation traits were calculated by summing available constituent glycoforms according to pre-specified formulas; traits for which none of the constituent glycoforms were detected were excluded from downstream analyses (See **Supplementary file 2.csv** which shows individual glycoforms summing or representing each trait).

### Statistical Analysis

All data processing and statistical analyses were conducted using JMP 18.2.1 (SAS Institute, Cary, NC) and R Statistical Software (R Studio version 2025.05.1). Only participants in the CR intervention group were included in the current analyses.

Prior to statistical modeling, all glycoform traits were assessed for normality using Shapiro–Wilk tests and Q-Q plots in JMP. Non-normally distributed traits were subjected to transformations, including log and Johnson family transformations, and the transformation resulting in the closest approximation to a normal distribution (based on p-values) was retained for subsequent analyses. Longitudinal changes in glycoform traits were evaluated using repeated measures linear mixed-effects models (lme4 and nlme packages in R), with participant as a random effect and baseline, age, and sex included as covariates. Models assessed changes from baseline at 12 months, and 24 months, with effect sizes quantified via Cohen’s *f²*. False discovery rate (FDR) adjustment was applied to account for multiple comparisons. For traits showing significant longitudinal changes, line graphs were generated to visualize glycoform trajectories across the three time points.

To investigate the relationships between AGP glycoform changes and metabolic biomarkers, repeated-measures correlation (rmcorr, R), moderation analyses and LASSO (Least Absolute Shrinkage and Selection Operator) regression model (glmmLasso for linear mixed models, R) analyses were performed. Repeated measures correlation analyses were done on all glycoforms and anthropometric, clinical, immune and other metabolic biomarkers in CALERIE2 (without FDR correction), but the lasso models were only evaluated for significant rmcorr associations between glycoform and outcomes. Moderation analyses were performed for glycoforms that showed significant correlations in rmcorr and outcomes (leptin, body weight and physical activity) that were selected *a priori* based on literature. Moderation analysis for outcomes (leptin, body weight) was done using percent CR as the independent variable, and with including and the interaction between glycoform and leptin as well as age and sex as covariates; analyses were not FDR corrected. For the lasso - bootstrap (n = 100) penalized linear mixed-effects models were fitted separately for each clinical outcome. Glycoforms were entered as penalized fixed effects; sex, age, and time (BL, 12mo and 24mo) were included as un-penalized fixed effects; and, a participant-specific random intercept was included to account for correlation among repeated measurements from the same participant. Bootstrap selection frequencies were calculated to determine how stably the identified relationships were observed, and the directional stability of the relationship was also verified. When data were missing (since several variables were only available at 12mo and 24mo), the model was fitted only for 12mo and 24mo and leaving out the BL time point.

## Results

### Participant Demographics

Of the 26 participants from the CALERIE subset evaluated in this study, there were 13 men and women, all of whom (**Table 1**) maintained ∼12% CR as part of this 2 y intervention. All participants lost weight between BL-12mo (-9.3 kg) and gained back some of the lost weight between 12-24 mo (+1.1 kg). Body weight and BMI results were analyzed and presented in a previous manuscript [20] and hence, while data are shown, these analyses were not repeated here. Both light and moderate intensity physical activity energy expenditures were reduced at 24mo compared to BL in this sub-sample.

**Table 1.** Descriptive Characteristics of Study Sample at Baseline, 12 months, and 24 months.

|  | **Baseline** | **12 Months** | **24 Months** | **BL vs 12mo** | **12mo vs 24mo** | **BL vs 24mo** |
|---|---|---|---|---|---|---|
| **Gender (n)** |  |  |  |  |  |  |
| **Male** | 13 | 13 | 13 |  |  |  |
| **Female** | 13 | 13 | 13 |  |  |  |
| **Age (years)** | 39.2 ± 6.8 | 40.2 ± 6.8 | 41.2 ± 6.8 |  |  |  |
| **Body Weight (kg)** | 77.0 ± 9.5 | 67.7 ± 9.4 | 68.8 ± 9.7 | *<0.001* | *0.011* | *<0.001* |
| **BMI (kg/m<sup>2</sup>)** | 25.9 ± 1.9 | 22.8 ± 1.8 | 23.1 ± 2. 0 | *<0.001* | *0.012* | *<0.001* |
| **CR (%)** | – | 16.8 ± 7.1 | 8.0 ± 8.0 | *<0.001* |  |  |
| **Light intensity EE (kcal/day)** | 1476.5 ± 244.5 | 1366.5 ± 246.2 | 1401.1 ± 274.7 | *<0.001* | 0.409 | *0.019* |
| **Mod intensity EE (kcal/day)** | 118.2 ± 111.8 | 80.6 ± 88.1 | 64.8 ± 59.4 | 0.137 | 0.693 | *0.021* |
| **Hard intensity EE (kcal/day)** | 22.3 ± 41.8 | 23.8 ± 31.2 | 45.4 ± 78.8 | 0.993 | 0.244 | 0.201 |
| **V. Hard intensity EE (kcal/day)** | 46.2 ± 94.4 | 71.0 ± 148.0 | 53.5 ± 162.8 | 0.656 | 0.809 | 0.964 |

Values are presented as mean ± SD. BMI = body mass index; EE = Energy Expenditure

### Longitudinal changes in AGP glycoforms

17 derived glycan traits showed significant longitudinal variation across baseline, 12-month, and 24-month timepoints (**Figure 1**). Most changes occurred by 12 months, followed by partial attenuation or stabilization by 24 months. Of the traits with complete pairwise contrasts, only three (site IV total fucosylation and site V tetra-sialylation and tri-antennary reduction, all AGP1) remained significantly different from baseline at both 12 and 24 months, while the majority (11 of 15) were significant at 12 months but no longer distinguishable from baseline by 24 months. One trait, site IV tetra-antennary glycans, showed the inverse pattern, reaching significance only at 24 months.

**Figure 1.**
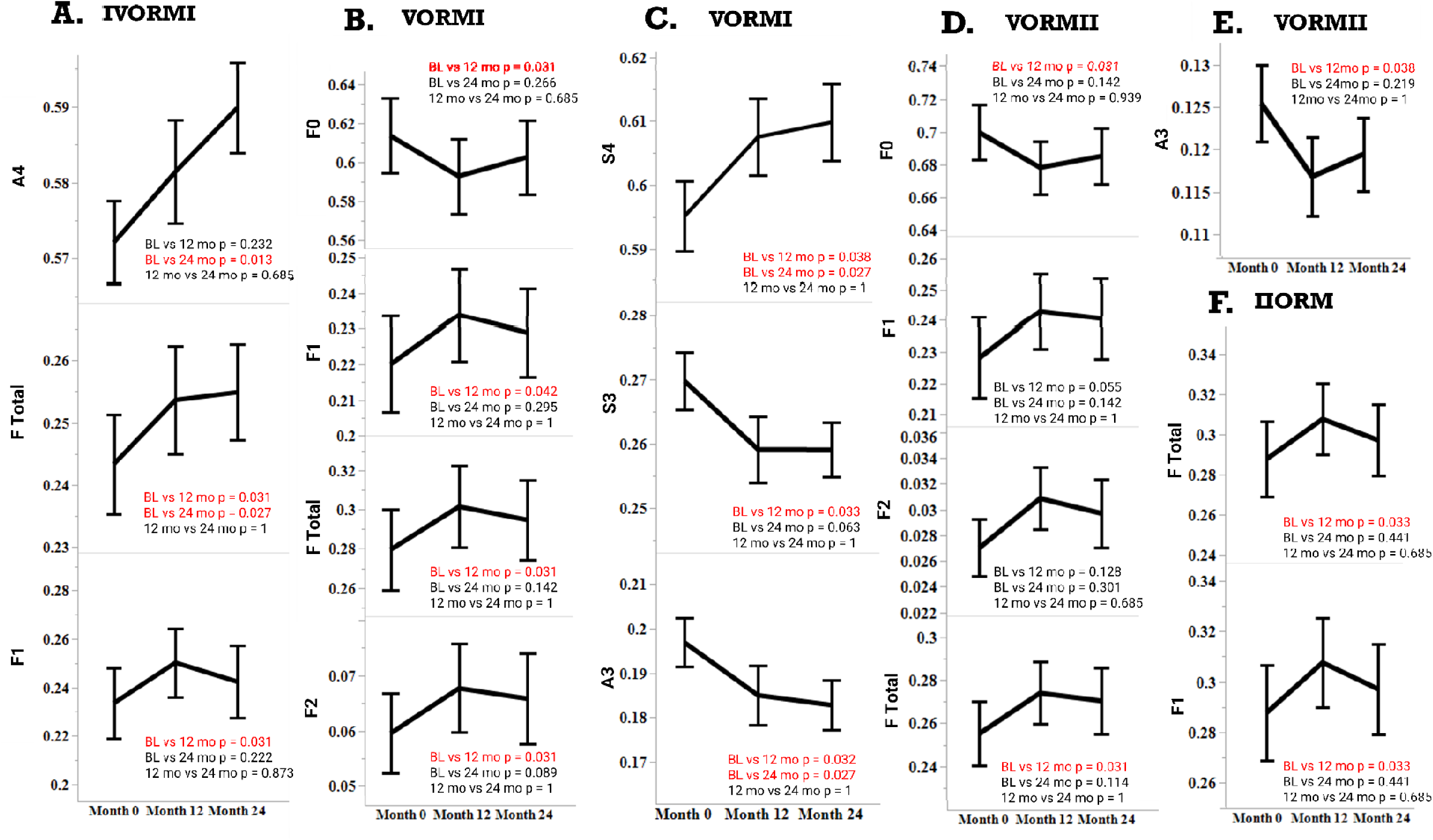
Longitudinal changes in α1-acid glycoprotein (AGP) glycosylation during calorie restriction. Relative abundances of AGP glycoform traits (A–F) in plasma measured at baseline, 12 months, and 24 months during the 2-year calorie restriction (CR) intervention (*n* = 26). Panels represent AGP glycoform traits corresponding to specific N-linked glycosylation sites and glycan compositions, derived from mass spectrometry–based glycoproteomics. Data were evaluated using linear mixed-effects models with visit month as a repeated measure, participant as a random effect, and baseline value, sex, and age included as covariates, followed by false discovery rate (FDR) correction. Significant longitudinal effects are highlighted in red. A – IV ORM-I, B &C – V-ORM-I, D&E – V-ORM-II, F-II-ORM.

Fucosylated glycans at glycosylation site V (Asn-103) of both AGP1 and 2 increased at both 12 and 24mo and were the most consistent and pronounced changes due to CR. Fucosylation changes at sites II (Asn56) and IV (Asn93) were also observed at 12mo, and continued to rise at site IV, but not in site II. Changes in branching of glycans on AGP were mixed – in AGP1 at site IV (Asn93), tetraantennary rose both at 12 and 24mo compared to 0mo, but in site V (Asn103) both in AGP1 and AGP2 triantennary glycans reduced at 12mo compared to 0mo. This reduction persisted at 24mo only in AGP1 site 5, and not in AGP2. Cohen’s *f²* values (0.10–0.25) suggested small-to-moderate effect sizes (**Supplementary table 2**). Model performance was strong across traits. These results indicate site-specific and structure-dependent modulation of AGP glycosylation over the intervention period.

### Glycoform associations with anthropometric, clinical and metabolic outcomes across time

Repeated-measures correlation analysis identified numerous associations between AGP glycan traits and measures of adiposity and metabolic function (**Supplementary figure 2**). Body weight and BMI had the greatest number of significant associations (∼42) with glycan traits – almost equally divided amongst positive and negative ones across various derived glycan feature configurations and sites. No distinct single glycan trait, site, or direction was observed. This was closely followed by triglycerides, 120-minute insulin, HOMA-β, leptin, and several C-peptide measures that had more than 10 significant associations. These associations were predominantly inverse. Insulin at 30 minutes and IGFBP-1 also showed 13 and 10 significant associations, whereas HOMA-β, 120-minute glucose, 30-minute glucose, and cholesterol showed fewer than 10 associations. The moderation analyses identified site I reduced branching/antennarity and site IV tetra-sialylation to influence the relationship between CR and leptin. (**Supplementary Table 3**).

The glmmLasso identified that most of the pairwise rmcorr associations did not translate into stable regression relationships (**Figure 2-A**). None of the body weight, BMI and glycan trait relationships were seen in the bootstrapped glmmLasso models. For triglycerides and leptin, all retained glycan traits had very low bootstrap selection frequencies, generally 1–2%, indicating weak and unstable multivariable signals. The 120-minute insulin models were numerically unstable (large coefficients and prediction errors) and were not interpreted. For 30-minute insulin, several glycoforms were retained, but selection frequencies remained low, similar to triglycerides and leptin. It is also important to note here that insulin (fasting and postprandial) was only available at 12mo and 24mo.

**Figure 2.**
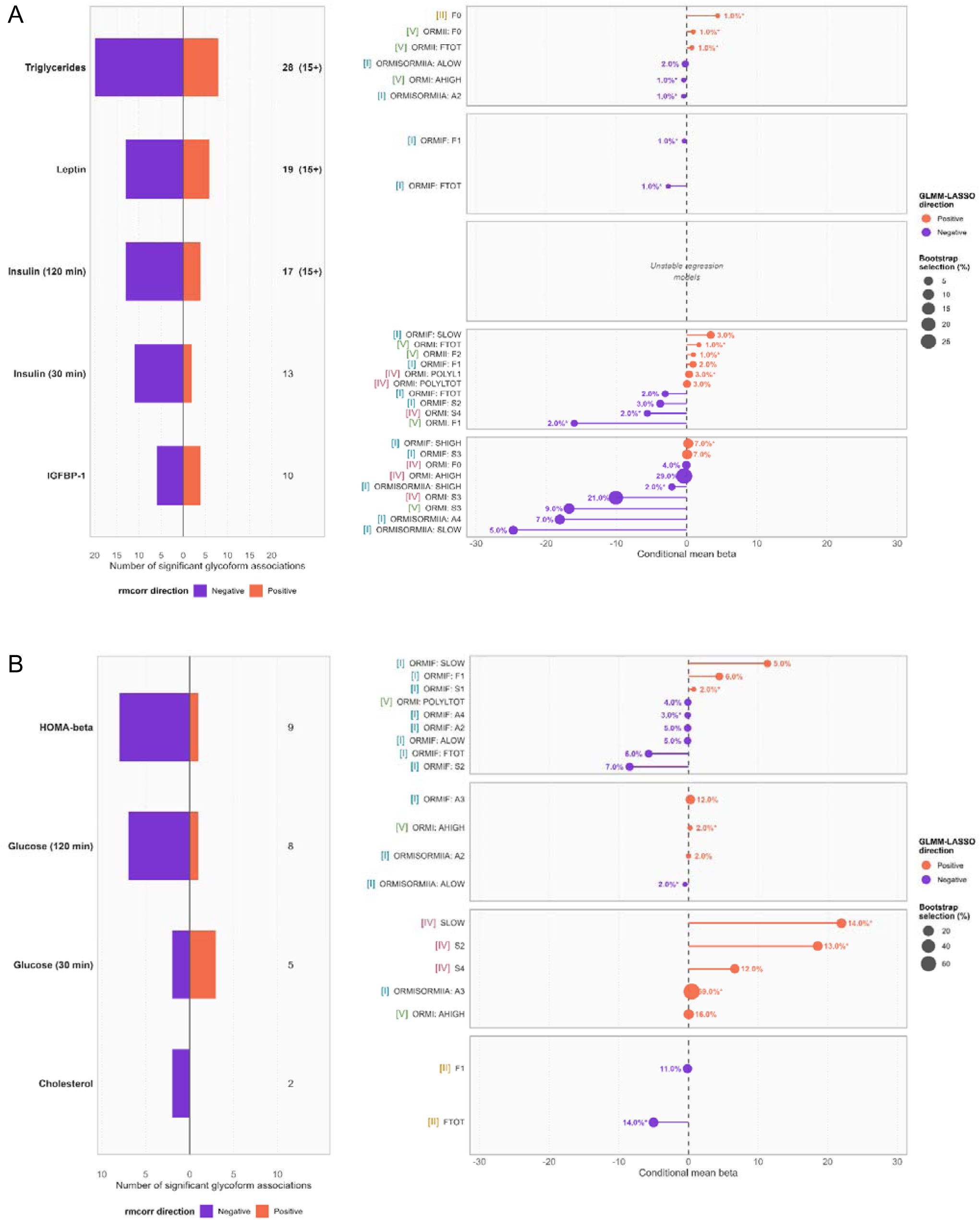
RMCORR screening and bootstrap GLMM-LASSO results for modeled clinical outcomes. Clinical and metabolic outcomes were screened using repeated-measures correlation (rmcorr), then modeled using bootstrap glmmLasso. Only outcomes which could fit glmmLasso models are plotted, so despite variables like clinical weight and BMI having 15+ rmcorr associations with glycoforms, they are not depicted because no valid model was fit. (A) Outcomes with 10 or more significant rmcorr associations and (B) outcomes with fewer than 10 significant rmcorr associations. Left subpanels show the number of significant rmcorr associations for each outcome, with purple indicating negative and orange indicating positive associations (*p* < 0.05, not FDR corrected). Right subpanels show dot-forest plots of glycoforms retained in bootstrap glmmLasso models, where the x-axis represents the conditional mean beta, dot color reflects coefficient direction, and dot size and adjacent labels reflect bootstrap selection frequency. An asterisk (*) denotes directionally consistent selection across bootstrap samples. Roman numerals indicate AGP/ORM glycosylation site (I–V). Outcomes labeled 15+ met the screening reference of at least 15 significant glycoform associations. Regression models for 120-minute insulin were unstable with large coefficients and prediction errors and were not interpreted. Several outcomes were only available at mo 12 and 24: n for each time point as BL (baseline) 12m (12 month) and 24m (24 month); Insulin fasting (BL: 0, 12m: 26, 24m: 26), Insulin 30 mins (BL: 0, 12m: 24, 24m: 26), Insulin 60 mins (BL: 0, 12m: 24, 24m: 26), Insulin 120 mins (BL: 0, 12m: 24, 24m: 26) and HOMA-beta (BL: 0, 12m: 26, 24m: 26) and Inflammatory markers: CRP (BL: 0, 12m:26, 24m:26), IL1B (BL: 0, 12m: 13, 24m: 0), IL6 (BL: 0, 12m:26, 24m: 25), and IL8 (BL:0, 12m: 26, and 24m:26). *Abbreviations: SLOW-sialylation low (0 or 1), SHIGH – sialylation high (*≥*2), AHIGH – high antennary/branching (>2, 3 or 4), ALOW – low antennary/branching (*≤*2), F – fucosylation (F1, F2 etc are mono, di fucosylation, FTOT-total fucosylation), S-sialylation (S1, S2 etc are mono, di sialylation), A – antennary (branching – A2, A3 are bi, tri antennary etc), polyL1 – poly N-acetyllactosamine. ORMI – AGP1, ORMII – AGP2, ORMIF – F variant of the same AGP family. ORMISORMIIA - peptide sequence shared between specific allelic variants of AGP1 and AGP2 (ORMIS and ORMIIA) that cannot be distinguished by the MS workflow and treated as a joint AGP1/AGP2 glycopeptide species*.

IGFBP-1 retained more associations with glycan traits, the strongest retained association being a negative coefficient for the site-IV A-high (tri or tetra-antennary) trait, selected in 29% of bootstrap samples, followed by a negative coefficient for site-IV S3 (tri-sialylation) trait selected in 21%. Several additional negatively associated traits were selected in 5–9% of bootstraps, whereas the positively associated traits were selected in only 7% of bootstraps. Important to note that IGFBP-1 was only available at 12mo and 24mo time points.

Among outcomes with fewer than 10 rmcorr associations (**Figure 2-B**), HOMA-β and 120-minute glucose generally showed low bootstrap selection frequencies. In contrast, 30-minute glucose had the most stable glmmLasso result at site-I A3 (triantennary) trait which was positively associated and selected in 69% of bootstrap samples. Additional positive associations at site-IV S-low, S2, and S4 were selected in 12–14% of samples. With cholesterol, site-II total fucosylation was negatively associated and selected in 14% of bootstrap samples, while site-II F1 was selected in 11%. All these variables were also only available at 12mo and 24mo time points.

## Discussion

Our pilot investigation suggests that CR induces site-specific remodeling of the AGP1/AGP2 N-glycome. Following 24 mo of CR, higher fucosylation was observed at all sites, along with greater tetra-antennary glycans and fewer tri-antennary glycans at sites IV and V, and greater tetra-sialylation at site V. In our exploratory correlation and penalized regression analyses, greater site-I tri-antennary glycan abundance co-varied with greater 30-minute glucose, found in 69% of bootstrap models for our most stable model. At site IV, greater branching (≥3) and tri-sialylation were associated with lower IGFBP-1 (selected in 29% and 21% of bootstrap models). A lower IGFBP-1 is generally consistent with greater insulin-mediated suppression of hepatic IGFBP-1[28], and commonly associated with higher insulin resistance[29,30]. However, considering that these metabolic measures were only available in the CR maintenance phase of the intervention (ie 12-24 mo), this does not reflect what happened at the onset of CR, which involved greater weight loss, and a ∼17% CR at BL – 12mo vs 8% in 12-24mo. Finally, specific glycoforms may moderate the effect of CR on leptin. However, these need further investigations to confirm and verify.

During CR, the increase in tetra-antennary glycans at site IV and reduction in tri-antennary glycans at site V are consistent with a reduced inflammatory physiological state [12,31]. Generally, half or even greater proportion of glycans at site IV and V are tetra-antennary. Notably, both in acute and chronic inflammation, glycans at these sites shift towards lower branching complexity[12,31]. In the current study, with CR, the changes observed in branching at these sites could be associated with reduction in inflammation, particularly between BL and 12mo, when most of these changes occurred. The increase in tetra-antennary glycans on site IV and reduction in tri-antennary glycans in site V are the only sustained changes from BL through 24mo, and that likely reflects the shift in systemic inflammation status with CR.

AGP1/2 glycans originating from the liver (which is the most predominant type of AGP in circulation) are generally highly fucosylated, and can be both antennary (α1,3 or α1,4) and core fucosylated (α 1,6), although the latter is noted to be a minor proportion of total fucosylation, or even absent [32-34]. Non-hepatic (seminal/prostatic) AGP has been reported to have more α1,4 fucosylated glycans, but this is not the predominant source of AGP in circulation[35]. In hepatic-origin AGP, α1,3 fucosylation is the most common fucosyl-link, particularly in sites II, III and IV, which are involved heavily in immunomodulation, and have been reported to be elevated in acute and chronic inflammatory conditions [12,36,37]. However, its ability to module inflammation is contingent on the presence of α2,3 sialyl groups on the same glycan chain and formation of sialyl-Lewis X structures[17]. While there was an increase in tetrasialylation at site V with a concomitant decrease in trisialylation, their configuration (ie α 2,3 or α 2,6) is not known. Also, the formation of sialyl Lewis X structures themselves are not necessarily pro-inflammatory, and as stated earlier, have been noted to be immunomodulatory. Hence, the link between this increase in fucosylation and inflammation remains unclear and speculative. Given our understanding and this newly generated data, it is evident that CR modulates AGP glycans to be more antennary, along with greater terminal sialic acid and fucose additions, however the net inflammatory effect of these changes remain unknown and warrants further investigation.

While AGP is an acute phase inflammatory protein, in recent years, it has become associated with leptin, food intake, energy balance and insulin resistance [8,38,39]. Specifically, AGP in circulation is positively correlated with leptin, insulin, body adiposity and insulin resistance in mouse and humans (but sex dependent in the latter) [8,39]. In mice, AGP1 can also bind to leptin receptor, activate JAK2-STAT3 signaling cascade, and reduce food intake and body weight [38], but these effects were not observed in humans[40]. So, while we did observe correlation relationships between leptin, food intake and some AGP glycoforms, none of them were sustained in our robust-glmmLasso models. Our exploratory moderation analyses identified lower antennarity at site I and higher sialylation at site IV to potentially influence the effect of CR on leptin, however, these remain hypotheses that need to be verified further. It is likely that AGP protein concentration (which we did not measure) plays a role in leptin, body weight/adipose, and insulin regulation. And, we did not have the power to identify if specific glycoforms are critical to this process.

In this pilot study, we observed a positive relationship between circulating glucose at 30 mins and site-I tri-antennary glycans, and an inverse relationship between IGFBP-1 and site-IV tri-and tetra-antennary glycans as well as trisialylation. These findings raise the possibility that the metabolic significance of AGP branching differs by glycosylation site. The positive association between site-I tri-antennary glycans and 30-minute glucose is consistent with what has been observed in past studies about links between greater branching of the total plasma N-glycome to adverse metabolic profiles. Conversely, the interpretation of increased branching at sites IV and V as consistent with reduced inflammation derived from prior site-specific AGP studies. These observations could suggest that assigning a uniform inflammatory or metabolic interpretation to AGP branching irrespective of site might be inaccurate. This site-level heterogeneity, or meta-heterogeneity, has been noted of various other proteins [41]. And future studies need to establish that sites I and IV/V in AGP1 do respond to distinct regulatory mechanisms, and further investigate what these mechanisms are.

## Data Availability

All data will be submitted to the NIA Aging Research Biobank and will be available upon reasonable request.

## Strengths, Limitations and Conclusions

The study used data and samples from CALERIE2 study, the longest, largest and most well-controlled CR intervention in non-obese adults till date. The inquiry is novel, and our analyses have developed several hypotheses that warrant further investigation. Since very little is known about the role of AGP glycoforms in CR induced metabolic changes in adults, and in numerous other physiological states related to immunomodulation, this study provides the first insight into this. However, the study sample size is small, albeit a pilot, and does not have a control group. Both of these limit the ability of the data to be used for generating robust evidence to test/support hypotheses.

