## Supplemental File for "Remodeling of α1-Acid Glycoprotein Glycoforms During Long-Term Calorie Restriction"

**Supplementary Table 1. Structural classification and nomenclature of α1-acid glycoprotein (AGP) N-glycopeptides analyzed in the study.**

| **Derived Glycan Trait** | **ORM Type** | **Glycosylation Stie** | **Description** |
|---|---|---|---|
| IORMIF__Alow. | I | I | Low branching |
| IORMIF__A2. | I | I | Bi-antennary |
| IIIORM__A3 | I or II | III | Tri-antennary |
| IORMIF__A4. | I | I | Tetra-antennary |
| IIIORM__Ahigh. | I or II | III | High branching |
| IORMIF__Slow. | I | I | Low-sialylation |
| IORMIF__S1. | I | I | Mono-sialylated |
| IORMIF__S2. | I | I | Di-sialylated |
| IORMIF__S3. | I | I | Tri-sialylated |
| VORMII__S4. | II | V | Tetra-sialylated |
| IORMIF__Shigh. | I | I | High-sialylation |
| IORMIF__F0. | I | I | Non-fucosylated |
| IORMIF__F1. | I | I | Mono-fucosylated |
| IORMISORMIIA__F2. | I & II | I | Di-fucosylated |
| VORMI__polyL1. | I | V | Single polylactosamine |
| VORMI__polyL2. | I | V | Double polylactosamine |

**Supplementary Table 1**: AGP glycoforms are organized by glycosylation site, and structural features including branching, sialylation, fucosylation, and polylactosamine extension.

**Abbreviations:** AGP = α1-acid glycoprotein; ORM = orosomucoid; SNFG = Symbol Nomenclature for Glycans.

**Notes:** Branching refers to the number of N-glycan antennas, with A2–A4 indicating bi-, tri-, and tetra-antennary structures, respectively. Ahigh denotes highly branched glycans (≥3 antennae), whereas Alow denotes low-branched structures (≤2 antennae). Sialylation is indicated as S1–S4, corresponding to the number of terminal sialic acid residues; Slow denotes monosialylated structures (S1), and Shigh denotes highly sialylated structures (S2–S4). Fucosylation status is denoted by F0 (non-fucosylated), F1 (mono-fucosylated), or F2 (di-fucosylated). Polylactosamine extension is indicated as polyL1 (single LacNAc repeat) or polyL2 (double LacNAc repeat).

**Supplementary Table 2: ANOVA model outcomes.**

| Glycoform | Month 0 | Month 12 | Month 24 | Δ 12mo | Δ 24mo | ANOVA Model Month | Sex | Age | Cohen's $f^2$ |
|---|---|---|---|---|---|---|---|---|---|
| **IVORMI__A4** | 0.572 ± 0.00316 | 0.581 ± 0.00316 | 0.590 ± 0.00316 | -0.00935 ± 0.00412 | -0.01777 ± 0.00412 | 0.0004 | 0.8639 | 0.3226 | 0.22 |
| **IVORMI__Ftot** | 0.243 ± 0.00209 | 0.254 ± 0.00209 | 0.255 ± 0.00209 | -0.01030 ± 0.00296 | -0.01154 ± 0.00296 | 0.0004 | 0.0052 | 0.1311 | 0.25 |
| **IVORMI__F1** | -0.3409 ± 0.0648 | -0.0549 ± 0.0648 | -0.1642 ± 0.0648 | -0.286 ± 0.0807 | -0.177 ± 0.0807 | 0.0034 | 0.271 | 0.0953 | 0.14 |
| **VORMI__F0** | 0.768 ± 0.0469 | 0.547 ± 0.0469 | 0.645 ± 0.0469 | 0.2206 ± 0.0592 | 0.1231 ± 0.0592 | 0.0021 | 0.0717 | 0.1571 | 0.15 |
| **VORMI__F1** | 0.220 ± 0.00348 | 0.234 ± 0.00348 | 0.229 ± 0.00348 | -0.01372 ± 0.00446 | -0.00883 ± 0.00446 | 0.0118 | 0.0656 | 0.1353 | 0.11 |
| **VORMI__Ftot** | -0.502 ± 0.0493 | -0.294 ± 0.0493 | -0.350 ± 0.0493 | -0.208 ± 0.0611 | -0.152 ± 0.0611 | 0.0039 | 0.1514 | 0.1752 | 0.13 |
| **VORMI__F2** | -1.43 ± 0.0507 | -1.22 ± 0.0507 | -1.26 ± 0.0507 | -0.2156 ± 0.0618 | -0.1725 ± 0.0618 | 0.0024 | 0.156 | 0.2262 | 0.14 |
| **VORMI__S4** | 0.595 ± 0.00301 | 0.608 ± 0.00301 | 0.610 ± 0.00301 | -0.01241 ± 0.00396 | -0.01469 ± 0.00396 | 0.0010 | 0.7425 | 0.1088 | 0.19 |
| **VORMI__S3** | 0.270 ± 0.00232 | 0.259 ± 0.00232 | 0.259 ± 0.00232 | 0.0106 ± 0.00322 | 0.0107 ± 0.00322 | 0.0016 | 0.9651 | 0.0926 | 0.2 |
| **VORMI__A3** | -1.63 ± 0.0158 | -1.70 ± 0.0158 | -1.71 ± 0.0158 | 0.0693 ± 0.0207 | 0.0762 ± 0.0207 | 0.0008 | 0.6968 | 0.1393 | 0.2 |
| **VORMII__F0** | 0.153 ± 0.0567 | -0.118 ± 0.0567 | -0.027 ± 0.0567 | 0.270 ± 0.0729 | 0.180 ± 0.0729 | 0.0019 | 0.107 | 0.5653 | 0.16 |
| **VORMII__F1** | 0.228 ± 0.0041 | 0.243 ± 0.0041 | 0.241 ± 0.0041 | -0.0150 ± 0.00508 | -0.0125 ± 0.00508 | 0.0104 | 0.106 | 0.6156 | 0.11 |
| **VORMII__F2** | -0.180 ± 0.069 | 0.177 ± 0.069 | -0.009 ± 0.069 | -0.356 ± 0.091 | -0.171 ± 0.091 | 0.0012 | 0.803 | 0.8144 | 0.18 |
| **VORMII__Ftot** | -0.155 ± 0.061 | 0.116 ± 0.061 | 0.055 ± 0.061 | -0.271 ± 0.079 | -0.210 ± 0.079 | 0.0031 | 0.1397 | 0.547 | 0.15 |
| **VORMII__A3** | 0.313 ± 0.094 | -0.092 ± 0.094 | 0.025 ± 0.094 | 0.404 ± 0.129 | 0.288 ± 0.129 | 0.0089 | 0.2993 | 0.0542 | 0.14 |
| **IIORM__Ftot** | 0.288 ± 0.0048 | 0.308 ± 0.0048 | 0.297 ± 0.0048 | -0.0198 ± 0.0061 | -0.0093 ± 0.0061 | 0.0084 | 0.1081 | 0.179 | 0.12 |
| **IIORM__F1** | 0.288 ± 0.0048 | 0.308 ± 0.0048 | 0.297 ± 0.0048 | -0.0198 ± 0.0061 | -0.0093 ± 0.0061 | 0.0084 | 0.1081 | 0.179 | 0.12 |

Supplementary Table 2: Cohens $f^2$ is interpreted as >0.02 = small effect size, >0.15 = medium effect size and > 0.35 = large effect size. $f^2$ is a ratio of $R^2$ values, and doesn't follow a simple distribution. No standard formula exists for its confidence intervals, so bootstrapping or variance propagation methods are usually used to estimate them. This makes $f^2$'s CI less stable than those for other effect sizes and were not calculated.

[Selya, A. S., Rose, J. S., Dierker, L. C., Hedeker, D. & Mermelstein, R. J. A Practical Guide to Calculating Cohen's f(2), a Measure of Local Effect Size, from PROC MIXED. *Front Psychol* **3**, 111 (2012). https://doi.org/10.3389/fpsyg.2012.00111]

**Supplementary Table 3:** The role of glycoforms in moderating the relationship between select metabolic variables (leptin and body weight, based on rmcorr observed associations) and calorie restriction. No FDR correction was performed because these are exploratory analyses, and considered hypotheses generating.

| Independent Variable | Dependent Variable | Time | Moderator | P-value (with age and sex adjusted) | P-value (without age and sex adjusted) |
|---|---|---|---|---|---|
| pctcr | LEPTIN | 12 Month | IIORM__F0 | 0.4743 | 0.4588 |
| pctcr | LEPTIN | 24 Month | IIORM__F0 | 0.7593 | 0.8999 |
| pctcr | LEPTIN | 12 Month | IORMIF__A2 | 0.8364 | 0.6726 |
| pctcr | LEPTIN | 24 Month | IORMIF__A2 | 0.6215 | 0.2726 |
| pctcr | LEPTIN | 12 Month | IORMIF__A4 | 0.484 | 0.464 |
| pctcr | LEPTIN | 24 Month | IORMIF__A4 | 0.605 | 0.7694 |
| pctcr | LEPTIN | 12 Month | IORMIF__Alow | 0.8364 | 0.6726 |
| pctcr | LEPTIN | 24 Month | IORMIF__Alow | 0.6215 | 0.2726 |
| pctcr | LEPTIN | 12 Month | IORMIF__F1 | 0.3979 | 0.3223 |
| pctcr | LEPTIN | 24 Month | IORMIF__F1 | 0.3392 | 0.7236 |
| pctcr | LEPTIN | 12 Month | IORMIF__Ftot | 0.3979 | 0.3223 |
| pctcr | LEPTIN | 24 Month | IORMIF__Ftot | 0.3392 | 0.7236 |
| pctcr | LEPTIN | 12 Month | IORMIF__S1 | 0.9615 | 0.9796 |
| pctcr | LEPTIN | 24 Month | IORMIF__S1 | 0.7728 | 0.5639 |
| pctcr | LEPTIN | 12 Month | IORMIF__S2 | 0.9557 | 0.9098 |
| pctcr | LEPTIN | 24 Month | IORMIF__S2 | 0.4258 | 0.1703 |
| pctcr | LEPTIN | 12 Month | IORMIF__Slow | 0.9605 | 0.9159 |
| pctcr | LEPTIN | 24 Month | IORMIF__Slow | 0.4349 | 0.1788 |
| pctcr | LEPTIN | 12 Month | IORMISORMIIA__A2 | 0.5541 | 0.2237 |
| pctcr | LEPTIN | 24 Month | IORMISORMIIA__A2 | *0.0465* | 0.1692 |
| pctcr | LEPTIN | 12 Month | IORMISORMIIA__Alow | 0.5541 | 0.2237 |
| pctcr | LEPTIN | 24 Month | IORMISORMIIA__Alow | *0.0465* | 0.1692 |
| pctcr | LEPTIN | 12 Month | IORMISORMIIA__S2 | 0.603 | 0.7736 |
| pctcr | LEPTIN | 24 Month | IORMISORMIIA__S2 | 0.6886 | 0.6298 |
| pctcr | LEPTIN | 12 Month | IVORMI__polyL1 | 0.2391 | *0.0474* |
| pctcr | LEPTIN | 24 Month | IVORMI__polyL1 | 0.794 | 0.3001 |
| pctcr | LEPTIN | 12 Month | IVORMI__polyLtot | 0.2391 | *0.0474* |
| pctcr | LEPTIN | 24 Month | IVORMI__polyLtot | 0.794 | 0.3001 |
| pctcr | LEPTIN | 12 Month | IVORMI__S4 | *0.0393* | *0.0076* |
| pctcr | LEPTIN | 24 Month | IVORMI__S4 | 0.9658 | 0.0954 |
| pctcr | LEPTIN | 12 Month | VORMI__F1 | 0.4021 | 0.4254 |
| pctcr | LEPTIN | 24 Month | VORMI__F1 | 0.7509 | 0.9289 |
| pctcr | LEPTIN | 12 Month | VORMI__polyL1 | 0.3691 | 0.4273 |
| pctcr | LEPTIN | 24 Month | VORMI__polyL1 | 0.692 | 0.3952 |
| pctcr | LEPTIN | 12 Month | VORMII__F0 | 0.8888 | 0.896 |
| pctcr | LEPTIN | 24 Month | VORMII__F0 | 0.6924 | 0.8901 |
| pctcr | LEPTIN | 12 Month | VORMII__F2 | 0.8606 | 0.9891 |
| pctcr | LEPTIN | 24 Month | VORMII__F2 | 0.6443 | 0.7846 |
| pctcr | CLINWT | 12 Month | IORMISORMIIA__S3 | 0.9266 | 0.7675 |
| pctcr | CLINWT | 24 Month | IORMISORMIIA__S3 | 0.6379 | 0.2815 |
| pctcr | LIGHTCAL | 12 Month | IORMISORMIIA__S3 | 0.5117 | 0.4598 |
| pctcr | LIGHTCAL | 24 Month | IORMISORMIIA__S3 | 0.9985 | 0.4846 |
| pctcr | BMI | 12 Month | IORMISORMIIA__Slow | 0.2458 | 0.2959 |
| pctcr | BMI | 24 Month | IORMISORMIIA__Slow | 0.9491 | 0.641 |
| pctcr | LIGHTCAL | 12 Month | VORMI__Shigh | 0.7231 | 0.5953 |
| pctcr | LIGHTCAL | 24 Month | VORMI__Shigh | 0.4108 | 0.9717 |
| pctcr | CLINWT | 12 Month | VORMI__Shigh | 0.5926 | 0.7817 |
| pctcr | CLINWT | 24 Month | VORMI__Shigh | 0.3209 | 0.749 |

**Supplementary Figure 1**

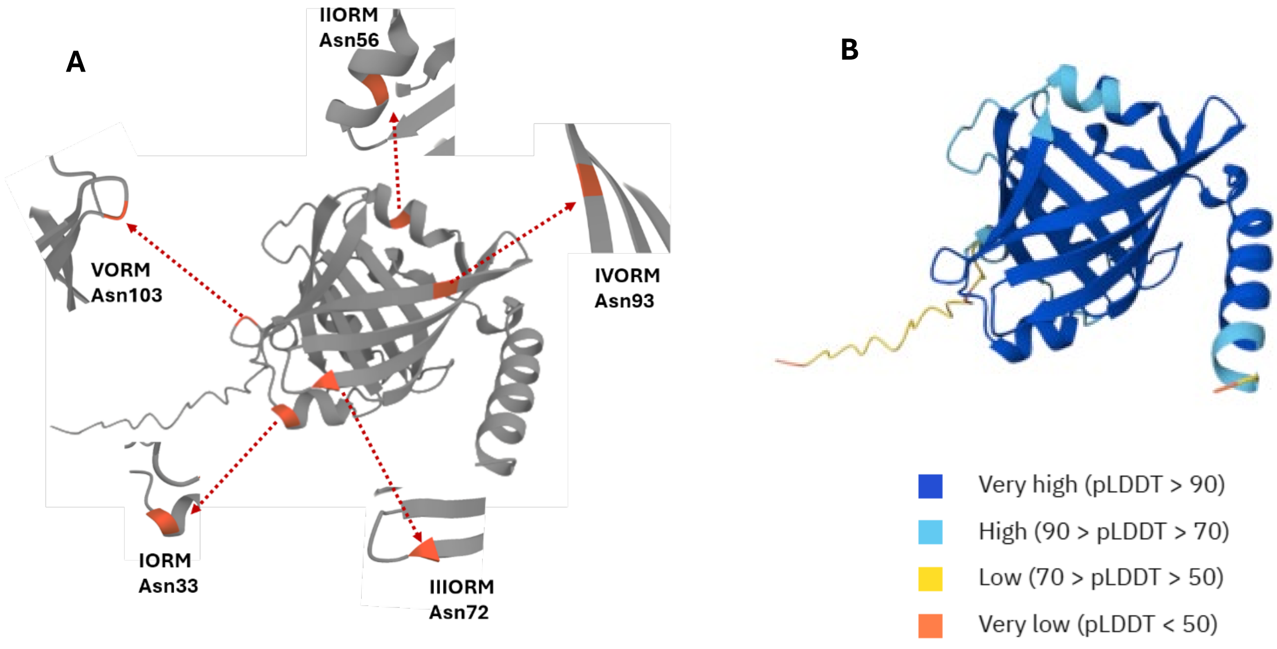


**Supplementary Figure 1**: The AGP1 protein (Homo sapiens) from AlphaFold (https://alphafold.ebi.ac.uk/entry/AF-P02763-F1). A - annotated with Asn-N-glycosylation sites, zoomed in sections shown radially. B – the degree of confidence for the structure denoted by pLDDT (the local Distance Difference Test) – the peer-residue estimate (superposition-free score) of its confidence on a scale of 0-100. Glycoform sites I, III, IV and V are modelled with high accuracy, whereas site II is modelled well. Overall, the N-glycosylation sites on AGP1 are modelled with acceptable confidence, and the detection of glycans at these sites are to be expected (both based on this and on literature).

**Supplementary Figure 2**

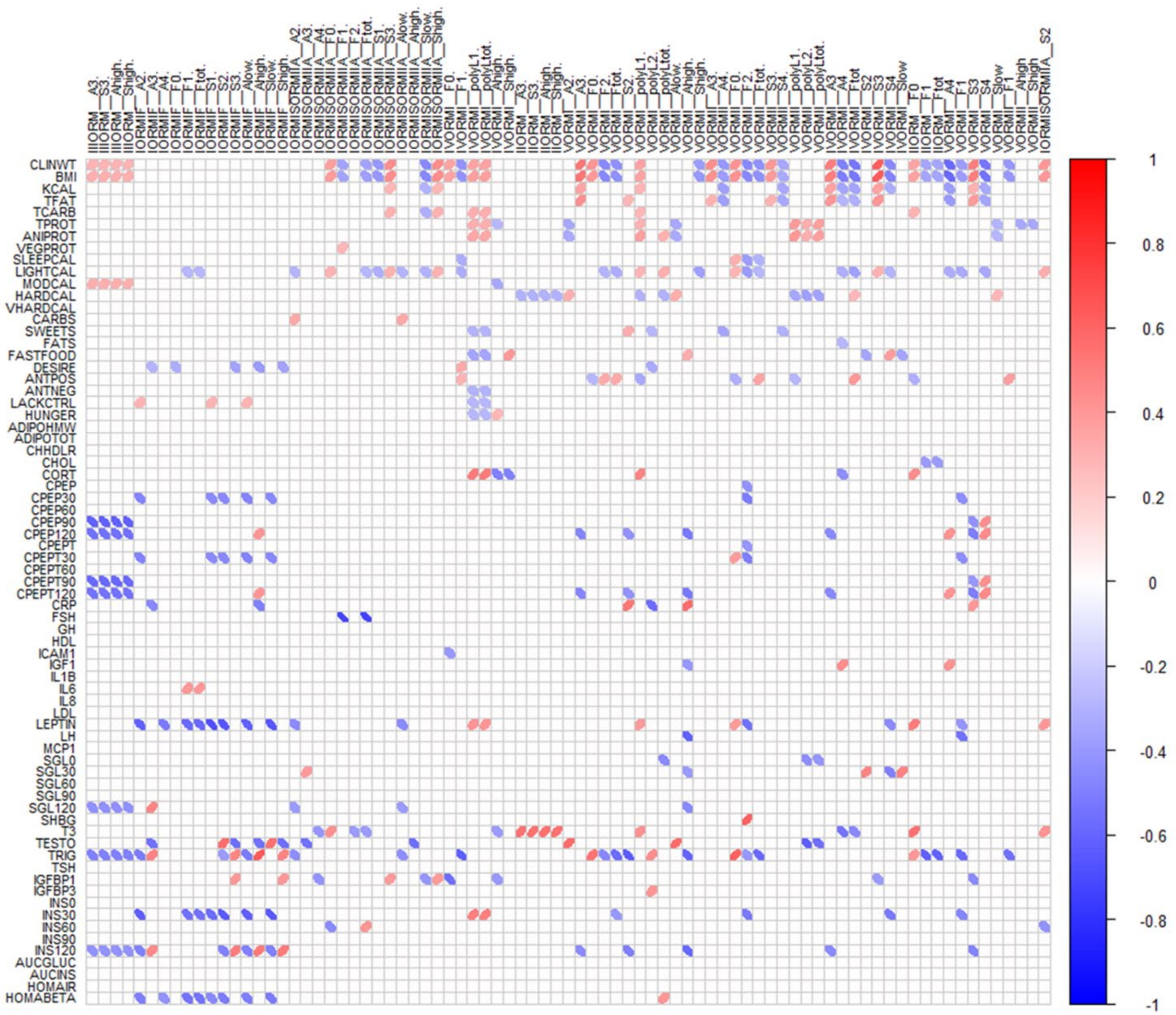


**Supplementary Figure 2**: Heat map of rmcorr p and r values for all variables and glycoforms. Blue ellipses indicate inverse relationships and red positive relationships; ellipse direction also displays positive or negative correlation. N for baseline, 12-month, and 24-month are 26 for all measures except * which denotes measures that were not measured at baseline (baseline n =0). ** denotes groups with missing measures as follows with n for each time point as BL (baseline) 12m (12 month) and 24m (24 month); Insulin fasting (BL: 0, 12m: 26, 24m: 26), Insulin 30 mins (BL: 0, 12m: 24, 24m: 26), Insulin 60 mins (BL: 0, 12m: 24, 24m: 26), Insulin 120 mins (BL: 0, 12m: 24, 24m:  26) and HOMA-beta (BL: 0, 12m: 26, 24m: 26) and Inflammatory markers: CRP (BL: 0, 12m:26,  24m:26), IL1B (BL: 0, 12m: 13, 24m: 0), IL6 (BL: 0, 12m:26, 24m: 25), and IL8 (BL:0, 12m: 26, and 24m:26).

**Supplementary File 2**

**Complete list of all derived apha-1-acid glycoprotein (AGP) N-glycosylation traits in this data.**

| Site Derived Trait | Formula* |
|---|---|
| IIIORM__A3 | IIIORM_N5H6S3 |
| IIIORM__S3 | IIIORM_N5H6S3 |
| IIIORM__Ahigh | IIIORM_N5H6S3 |
| IIIORM__Shigh | IIIORM_N5H6S3 |
| IORMIF__A2 | IORMIF_N4H5S1 + IORMIF_N4H5S2 |
| IORMIF__A3 | IORMIF_N5H6S2 + IORMIF_N5H6S3 + IORMIF_N5H6S2F1 + IORMIF_N5H6S3F1 |
| IORMIF__A4 | IORMIF_N6H7S3 + IORMIF_N6H7S3F1 |
| IORMIF__F0 | IORMIF_N4H5S1 + IORMIF_N4H5S2 + IORMIF_N5H6S2 + IORMIF_N5H6S3 + IORMIF_N6H7S3 |
| IORMIF__F1 | IORMIF_N5H6S2F1 + IORMIF_N5H6S3F1 + IORMIF_N6H7S3F1 |
| IORMIF__Ftot | IORMIF_N5H6S2F1 + IORMIF_N5H6S3F1 + IORMIF_N6H7S3F1 |
| IORMIF__S1 | IORMIF_N4H5S1 |
| IORMIF__S2 | IORMIF_N4H5S2 + IORMIF_N5H6S2 + IORMIF_N5H6S2F1 |
| IORMIF__S3 | IORMIF_N5H6S3 + IORMIF_N5H6S3F1 + IORMIF_N6H7S3 + IORMIF_N6H7S3F1 |
| IORMIF__Alow | IORMIF_N4H5S1 + IORMIF_N4H5S2 |
| IORMIF__Ahigh | IORMIF_N5H6S2 + IORMIF_N5H6S3 + IORMIF_N5H6S2F1 + IORMIF_N5H6S3F1 + IORMIF_N6H7S3 + IORMIF_N6H7S3F1 |
| IORMIF__Slow | IORMIF_N4H5S1 + IORMIF_N4H5S2 + IORMIF_N5H6S2 + IORMIF_N5H6S2F1 |
| IORMIF__Shigh | IORMIF_N5H6S3 + IORMIF_N5H6S3F1 + IORMIF_N6H7S3 + IORMIF_N6H7S3F1 |
| IORMISORMIIA__A2 | IORMISORMIIA_N4H5S1 + IORMISORMIIA_N4H5S2 |
| IORMISORMIIA__A3 | IORMISORMIIA_N5H6S2 + IORMISORMIIA_N5H6S3 + IORMISORMIIA_N5H6S2F1 + IORMISORMIIA_N5H6S3F1 |
| IORMISORMIIA__A4 | IORMISORMIIA_N6H7S3F1 + IORMISORMIIA_N6H7S3F2 |
| IORMISORMIIA__F0 | IORMISORMIIA_N4H5S1 + IORMISORMIIA_N4H5S2 + IORMISORMIIA_N5H6S2 + IORMISORMIIA_N5H6S3 |
| IORMISORMIIA__F1 | IORMISORMIIA_N5H6S2F1 + IORMISORMIIA_N5H6S3F1 + IORMISORMIIA_N6H7S3F1 |

| | |
|---|---|
| IORMISORMIIA__F2 | IORMISORMIIA_N6H7S3F2 |
| IORMISORMIIA__Ftot | IORMISORMIIA_N5H6S2F1 + IORMISORMIIA_N5H6S3F1 + IORMISORMIIA_N6H7S3F1 + IORMISORMIIA_N6H7S3F2 |
| IORMISORMIIA__S1 | IORMISORMIIA_N4H5S1 |
| IORMISORMIIA__S2 | IORMISORMIIA_N4H5S2 + IORMISORMIIA_N5H6S2 + IORMISORMIIA_N5H6S2F1 |
| IORMISORMIIA__S3 | IORMISORMIIA_N5H6S3 + IORMISORMIIA_N5H6S3F1 + IORMISORMIIA_N6H7S3F1 + IORMISORMIIA_N6H7S3F2 |
| IORMISORMIIA__Alow | IORMISORMIIA_N4H5S1 + IORMISORMIIA_N4H5S2 |
| IORMISORMIIA__Ahigh | IORMISORMIIA_N5H6S2 + IORMISORMIIA_N5H6S3 + IORMISORMIIA_N5H6S2F1 + IORMISORMIIA_N5H6S3F1 IORMISORMIIA_N6H7S3F1 + IORMISORMIIA_N6H7S3F2 |
| IORMISORMIIA__Slow | IORMISORMIIA_N4H5S1 + IORMISORMIIA_N4H5S2 + IORMISORMIIA_N5H6S2 + IORMISORMIIA_N5H6S2F1 |
| IORMISORMIIA__Shigh | IORMISORMIIA_N5H6S3 + IORMISORMIIA_N5H6S3F1 + IORMISORMIIA_N6H7S3F1 + IORMISORMIIA_N6H7S3F2 |
| IVORMI__A3 | IVORMI_N5H6S2 + IVORMI_N5H6S3 +IVORMI_N5H6S3F1 |
| IVORMI__A4 | IVORMI_N6H7S2 + IVORMI_N6H7S3 + IVORMI_N6H7S4 + IVORMI_N6H7S2F1 + IVORMI_N6H7S3F1 + IVORMI_N6H7S4F1 + IVORMI_N7H8S3 |
| IVORMI__F0 | IVORMI_N5H6S2 + IVORMI_N5H6S3 + IVORMI_N6H7S2 + IVORMI_N6H7S3 + IVORMI_N6H7S4 + IVORMI_N7H8S3 |
| IVORMI__F1 | IVORMI_N5H6S3F1 + IVORMI_N6H7S2F1 + IVORMI_N6H7S3F1 + IVORMI_N6H7S4F1 |
| IVORMI__Ftot | IVORMI_N5H6S3F1 + IVORMI_N6H7S2F1 + IVORMI_N6H7S3F1 + IVORMI_N6H7S4F1 |
| IVORMI__S1 | IVORMI_N6H7S1 |
| IVORMI__S2 | IVORMI_N5H6S2 + IVORMI_N6H7S2 + IVORMI_N6H7S2F1 |
| IVORMI__S3 | IVORMI_N5H6S3 + IVORMI_N5H6S3F1 + IVORMI_N6H7S3 + IVORMI_N6H7S3F1 + IVORMI_N7H8S3 |
| IVORMI__S4 | IVORMI_N6H7S4 + IVORMI_N6H7S4F1 |
| IVORMI__polyL1 | IVORMI_N7H8S3 |
| IVORMI__polyLtot | IVORMI_N7H8S3 |
| IVORMI__Ahigh | IVORMI_N5H6S2 + IVORMI_N5H6S3 + IVORMI_N5H6S3F1 + IVORMI_N6H7S2 + IVORMI_N6H7S3 + IVORMI_N6H7S4 + |

| | |
|---|---|
| | IVORMI_N6H7S2F1 + IVORMI_N6H7S3F1 + IVORMI_N6H7S4F1 + IVORMI_N7H8S3 |
| IVORMI__Slow | IVORMI_N5H6S2 + IVORMI_N6H7S2 + IVORMI_N6H7S2F1 |
| IVORMI__Shigh | IVORMI_N5H6S3 + IVORMI_N5H6S3F1 + IVORMI_N6H7S3 + IVORMI_N6H7S4 + IVORMI_N6H7S3F1 + IVORMI_N6H7S4F1 + IVORMI_N7H8S3 |
| IIORM__A3 | IIORM_N5H6S3 + IIORM_N5H6S3F1 |
| IIORM__F0 | IIORM_N5H6S3 |
| IIORM__F1 | IIORM_N5H6S3F1 |
| IIORM__Ftot | IIORM_N5H6S3F1 |
| IIORM__S3 | IIORM_N5H6S3 + IIORM_N5H6S3F1 |
| IIORM__Ahigh | IIORM_N5H6S3 + IIORM_N5H6S3F1 |
| IIORM__Shigh | IIORM_N5H6S3 + IIORM_N5H6S3F1 |
| VORMI__A2 | VORMI_N4H5S2 |
| VORMI__A3 | VORMI_N5H6S2 + VORMI_N5H6S3 + VORMI_N5H6S3F1 |
| VORMI__A4 | VORMI_N6H7S3 + VORMI_N6H7S4 + VORMI_N6H7S3F1 + VORMI_N6H7S4F1 + VORMI_N6H7S4F2 + VORMI_N7H8S3 + VORMI_N7H8S4 + VORMI_N8H9S3 |
| VORMI__F0 | VORMI_N4H5S2 + VORMI_N5H6S2 + VORMI_N5H6S3 + VORMI_N6H7S3 + VORMI_N6H7S4 + VORMI_N7H8S3 + VORMI_N7H8S4 + VORMI_N8H9S3 |
| VORMI__F1 | VORMI_N5H6S3F1 + VORMI_N6H7S3F1 + VORMI_N6H7S4F1 |
| VORMI__F2 | VORMI_N6H7S4F2 |
| VORMI__Ftot | VORMI_N5H6S3F1 + VORMI_N6H7S3F1 + VORMI_N6H7S4F1 + VORMI_N6H7S4F2 |
| VORMI__S2 | VORMI_N4H5S2 + VORMI_N5H6S2 + |
| VORMI__S3 | VORMI_N5H6S3 + VORMI_N5H6S3F1 + VORMI_N6H7S3 + VORMI_N6H7S3F1 + VORMI_N7H8S3 + VORMI_N8H9S3 |
| VORMI__S4 | VORMI_N6H7S4 + VORMI_N6H7S4F1 + VORMI_N6H7S4F2 + VORMI_N7H8S4 |
| VORMI__polyL1 | VORMI_N7H8S3 + VORMI_N7H8S4 |
| VORMI__polyL2 | VORMI_N8H9S3 |
| VORMI__polyLtot | VORMI_N7H8S3 + VORMI_N7H8S4 + VORMI_N8H9S3 |
| VORMI__Alow | VORMI_N4H5S2 |
| VORMI__Ahigh | VORMI_N5H6S2 + VORMI_N5H6S3 + VORMI_N5H6S3F1 + VORMI_N6H7S3 + VORMI_N6H7S4 + VORMI_N6H7S2F1 + |

| | |
|---|---|
| | VORMI_N6H7S3F1 + VORMI_N6H7S4F1 + VORMI_N6H7S4F2 + VORMI_N7H8S3 + VORMI_N7H8S4 + VORMI_N8H9S3 |
| VORMI__Slow | VORMI_N4H5S2 + VORMI_N5H6S2 |
| VORMI__Shigh | VORMI_N5H6S3 + VORMI_N5H6S3F1 VORMI_N6H7S3 + VORMI_N6H7S4 + VORMI_N6H7S3F1 + VORMI_N6H7S4F1 + VORMI_N6H7S4F2 + VORMI_N7H8S3 + VORMI_N7H8S4 + VORMI_N8H9S3 |
| VORMII__A3 | VORMII_N5H6S3 |
| VORMII__A4 | VORMII_N6H7S3 + VORMII_N6H7S4 + VORMII_N6H7S4F1 + VORMII_N6H7S4F2 + VORMII_N7H8S4 + VORMII_N7H8S4F1 + VORMII_N8H9S4 |
| VORMII__F0 | VORMII_N5H6S3 + VORMII_N6H7S3 + VORMII_N6H7S4 + VORMII_N7H8S4 + VORMII_N8H9S4 |
| VORMII__F1 | VORMII_N6H7S4F1 + VORMII_N7H8S4F1 |
| VORMII__F2 | VORMII_N6H7S4F2 |
| VORMII__Ftot | VORMII_N6H7S4F1 + VORMII_N6H7S4F2 + VORMII_N7H8S4F1 |
| VORMII__S3 | VORMII_N5H6S3 + VORMII_N6H7S3 |
| VORMII__S4 | VORMII_N6H7S4 + VORMII_N6H7S4F1 + VORMII_N6H7S4F2 + VORMII_N7H8S4 + VORMII_N7H8S4F1 + VORMII_N8H9S4 |
| VORMII__polyL1 | VORMII_N7H8S4 + VORMII_N7H8S4F1 |
| VORMII__polyL2 | VORMII_N8H9S4 |
| VORMII__polyLtot | VORMII_N7H8S4 + VORMII_N7H8S4F1 + VORMII_N8H9S4 |
| VORMII__Ahigh | VORMII_N5H6S3 + VORMII_N6H7S3 + VORMII_N6H7S4 +VORMII_N6H7S4F1 + VORMII_N6H7S4F2 + VORMII_N7H8S4 + VORMII_N7H8S4F1 + VORMII_N8H9S4 |
| VORMII__Shigh | VORMII_N5H6S3 + VORMII_N6H7S3 + VORMII_N6H7S4 + VORMII_N6H7S4F1 + VORMII_N6H7S4F2 + VORMII_N7H8S4 + VORMII_N7H8S4F1 + VORMII_N8H9S4 |
